# Clonal hematopoiesis of indeterminate potential-associated non-small cell lung cancer risk is potentiated by small particulate matter air pollution among non-smokers: a novel somatic variant–environment interaction

**DOI:** 10.1101/2024.01.17.24301439

**Authors:** Caitlyn Vlasschaert, Marco Buttigieg, Yash Pershad, Matthew Lanktree, Melinda C. Aldrich, Michael J. Rauh, Alexander G. Bick

## Abstract

Small particulate matter air pollution (PM_2.5_) is a recognized driver of non-small cell lung cancer (NSCLC) among non-smoking individuals. Inhaled PM_2.5_ recruits pro-inflammatory macrophages to the air-lung interface, which promotes malignant lung epithelial cell growth and progression to overt cancer. We sought to determine whether clonal hematopoiesis of indeterminate potential (CHIP), a common age-related condition characterized by hyperinflammatory macrophages, exacerbates PM_2.5_-associated NSCLC in non-smokers using genetic, environmental, and phenotypic data from 413,901 individuals in the UK Biobank. Among non-smokers, PM_2.5_ is not associated with NSCLC and not associated with prevalence of CHIP, but CHIP is associated with a doubling of NSCLC risk (hazard ratio (HR) 2.01, 95% confidence interval (CI): 1.34-3.00). Moreover, CHIP-associated NSCLC risk is exacerbated in the setting of above-median PM_2.5_ levels (HR 2.70, 95% CI: 1.60–4.55). PM_2.5_ × CHIP is also associated with significantly greater markers of systemic inflammation (CRP, IL-6, and IL-1β) than expected. Altogether, these results suggest CHIP and PM_2.5_ form a novel gene × environment interaction promoting NSCLC tumorigenesis in non-smokers.

## Introduction

Air pollution is a recently recognized driver of non-small cell lung cancers (NSCLCs), particularly among individuals who have never smoked.^1–4^ Mechanistically, inhaled small environmental particulate matter (≤ 2.5 µm diameter) recruits pro-inflammatory macrophages to the lungs. The proinflammatory microenvironment promotes clonal outgrowth of lung alveolar type II epithelial cells with pre-existing cancer driver mutations and progression to overt lung adenocarcinoma.^4^ Pro-inflammatory macrophages have been posited to interact with environmental factors to drive cancer progression in other settings, for example with asbestos leading to mesothelioma.^5^ Although many non-smokers are exposed to high levels of air pollution, relatively few develop lung cancer, suggesting that individual-level factors modify the risk of lung cancer in this setting. To date, no such factors have been identified.

Clonal hematopoiesis of indeterminate potential (CHIP) is a common age-related condition caused by mutations in hematopoietic stem cells (HSCs)^6^ that produce pro-inflammatory macrophages.^7^ CHIP has been associated with a greater prevalence and incidence of multiple chronic diseases^8–11^ and acute adverse events such as myocardial infarction and acute kidney injury^8,12^, seemingly due to inflammatory destruction of tissues and organs.^13–16^ CHIP has also been associated with an array of incident cancers and adverse outcomes among cancer patients^17–20^, though the risk varies heavily by cancer type, with one study even suggesting a protective effect for CHIP in colorectal cancer.^21^ Notably, an association between CHIP and incident lung cancer exists in current-, past-, and never-smoking populations, and a causal role for this relationship is supported by Mendelian randomization analyses.^18,20^ However, the mechanisms driving CHIP-associated lung cancer development in these populations are not known.

In this study, we evaluate the interaction between environmental particulate matter with aerodynamic diameter ≤ 2.5 µm (PM_2.5_) and CHIP in the development of incident non-small cell lung cancers (NSCLC). We find that CHIP but not PM_2.5_ is independently associated with incident NSCLC among non-smokers, but that the association between CHIP and NSCLC increases with higher PM_2.5_ exposure levels. This suggests that PM_2.5_ promotes NSCLC tumorigenesis in the setting of CHIP in non-smokers.

## Methods

### Exposure and outcome definitions

Data from all participants in the UK Biobank in whom CHIP was previously ascertained using whole exome sequencing (N = 451,095) was used for this study.^22^ CHIP was defined as a binary phenotype (present/absent) using a variant allele fraction (VAF) threshold of 2%, per its guideline definition.^23^ Additionally, the following subtypes of CHIP were defined: large CHIP clones (VAF ≥ 10%), and *DNMT3A*-CHIP and non-*DNMT3A* CHIP (i.e., CHIP where the driver mutation is in a gene other than *DNMT3A*). We examine the latter two subgroups because we and others have previously reported a greater effect for non-*DNMT3A* CHIP compared to *DNMT3A*-CHIP for various outcomes.^8,24,25^

Lung cancer events were ascertained using the cancer register data field codes 40006 (Type of cancer: ICD10), 40008 (Type of cancer: ICD9), 40011 (Histology of cancer tumour) and 40005 (Date of cancer diagnosis). Non-small cell lung cancer was defined as any lung cancer with “Histology of cancer tumour” other than small cell lung cancer. The follow-up period was from enrollment (2008-2012) until July 2021 (median 12.4 [interquartile range: 11.6–13.1] years). Particulate matter air pollution levels were derived from field codes 24006 (Particulate matter air pollution (pm2.5); 2010) and 24008 (Particulate matter air pollution 2.5-10um; 2010). These measurements reflect the average ambient particulate matter estimates for the year 2010, calculated by the UK Biobank using land use regression models.^26^ These data were unavailable for participants from Northern England and Scotland, and thus these participants in these geographic regions were excluded from our analyses.^27^ Particulate matter air pollution levels were examined as a continuous variable and as a dichotomous variable (*i*.*e*., above-median vs. below-median).

Smoking behavior was defined using field codes 20116 (smoking status) and 20160 (ever smoked), where individuals were classified as never-smokers if they responded no to both questions.

Baseline serum C-reactive peptide levels were measured in all UK Biobank participants, and levels of two other circulating inflammatory proteins (IL-6 and IL-1β) were measured using OLINK proteomics in a subset of participants (N = 44,625).

### Statistics

Cox proportional hazards models were utilized to evaluate associations between incident NSCLC, and CHIP and particulate matter pollution levels. Interaction between CHIP and particulate matter air pollution were assessed. Age, age^2^, sex, body-mass index (BMI), annual household income before taxes (dichotomized as ≥ or < £31,000), and educational attainment (dichotomized into two categories: “degree or professional education” or “other level of educational attainment”), and 10 principal components (PCs) of genetic ancestry were used as covariates. The choice of covariates was informed by prior studies of air pollution and lung cancer incidence in the UK Biobank.^4,27^ Cox proportional hazards analyses were run using the *survival* R package (version 3.5-7) and results are illustrated using the *metafor* (version 3.4-0) and *ggplot2* packages.

Logistic regression was used to evaluate the association between CHIP and PM_2.5_ levels. The associations between CHIP, PM_2.5,_ and inflammatory circulating protein levels were assessed using multiple linear regression including an interaction term for CHIP × PM_2.5_. As with the Cox proportional hazards analyses, these regression models were adjusted for age, age^2^, sex, BMI, household income, educational attainment, and 10 PCs of genetic ancestry. The OLINK proteomics-derived inflammatory marker levels were additionally adjusted for UK Biobank center and assay plate, per standard processing recommendations by the UK Biobank team.^28^ The map of air pollution estimates per-country shown in Supplemental Figure 1 was created using the 2019 air quality reports published by the WHO and visualized using the R packages *maps* (version 3.4.2) and *ggplot2* (version 3.4.4).

### Study approval

Access to the UK Biobank dataset was provided under applications 43397 (Vanderbilt University Medical Center) and 101583 (Queen’s University), and local approval for secondary analyses of the data was obtained from the Vanderbilt University Medical Center and Queen’s University institutional review boards.

## Results

Among 451,095 individuals in the prospective UK Biobank, 15,633 (3.4%) had CHIP and 247,560 (55%) were never-smokers. Of these, 413,901 individuals had air particulate matter levels measured, and the median small particulate matter air pollution level (PM_2.5_) was 9.93 [interquartile range (IQR): 9.28–10.56]. Relevant baseline characteristics for the study cohort are shown in Table 1.

**Table 1.** Baseline characteristics.

|  | **Current or past smokers**<br>(N = 186,317) | **Never smokers**<br>(N = 227,133) |
| --- | --- | --- |
| CHIP (N, %) | 7404 (4.0%) | 6916 (3.0%) |
| Age (years; mean ± SD) | 57.4 ± 7.9 | 55.9 ± 8.1 |
| Sex (% female) | 48.2 | 59.1 |
| Level of air particulate matter pollution<br>with diameter $\leq 2.5 \mu\text{m}$<br>($\mu\text{g}/\text{m}^3$; mean ± SD) | 10.05 ± 1.08 | 9.93 ± 1.03 |
| Level of air particulate matter pollution<br>with diameter 2.5-10 $\mu\text{m}$<br>($\mu\text{g}/\text{m}^3$; mean ± SD) | 6.44 ± 0.90 | 6.42 ± 0.90 |

There were 1983 cases of incident NSCLC in our cohort (1676 in current or past smokers and 307 in never-smokers) over a median of 12.5 years of follow-up. 137 of the 1676 current or past smokers who developed NSCLC (8%) had CHIP at study baseline, and 25 of the 307 (8.2%) never-smokers who developed NSCLC had CHIP at baseline. CHIP status was associated with incident NSCLC, as well as in current/past smokers and in never-smokers in analyses stratified by smoking status (**Figure 1A**; HR 1.73, 95% CI: 1.48–2.02, p = 3.7 x 10^−12^ in the analysis adjusted for smoking; HR 2.01, 95% CI: 1.34-3.00, p = 0.0007 in never-smokers). The point estimates among non-smokers were larger for large CHIP clones (HR 2.06, 95% CI: 1.28–3.33, p = 0.003) compared with small clones (HR 1.78, 95% CI: 0.88–3.59, p = 0.11) and similar for *DNMT3A*-CHIP and non-*DNMT3A* CHIP (HR 1.94, 95% CI: 1.17–3.21, p = 0.01 and HR 2.00, 95% CI: 1.06–3.75, p = 0.03, respectively).

**Figure 1.**
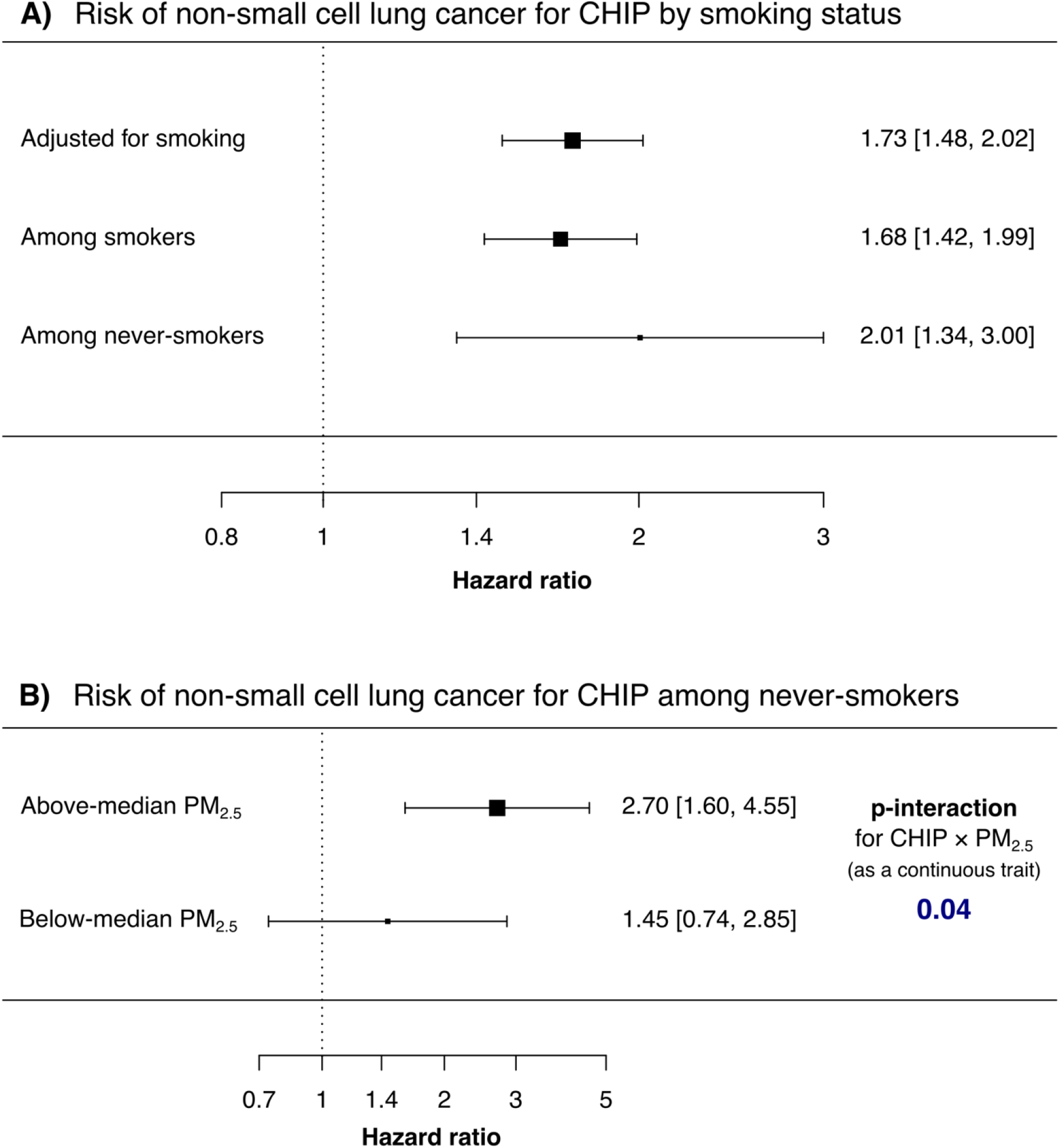
A) CHIP is associated with incident NSCLC irrespective of smoking status. B) CHIP is associated with NSCLC among never-smokers exposed to higher PM_2.5_ levels, and there is a significant interaction between CHIP and PM_2.5_ level.

Small particulate matter air pollution levels (i.e., PM_2.5_) were associated with incident NSCLC in current/past smokers (HR 1.17, 95% CI: 1.12–1.22 per µg/m^3^ increase in PM_2.5_ level), but not in never-smokers with or without CHIP added as a covariable (HR 1.00, 95% CI: 0.89–1.12 per µg/m^3^ increase in PM_2.5_ level in non-smokers in both models). However, in non-smokers there was a significant interaction between CHIP and PM_2.5_ levels (**Figure 1B**; HR 1.46, 95% CI: 1.02–2.11, p-interaction = 0.04). In the model with the interaction term, the association between CHIP and incident NSCLC was non-significant (HR 0.05, 95% CI: 0.001–1.94; p = 0.11), indicating that the CHIP-NSCLC risk was highly dependent upon PM_2.5_ levels in non-smokers. To further illustrate the interplay between CHIP and PM_2.5_ levels, we tested the association between CHIP and incident NSCLC in analyses stratified by median PM_2.5_. We observe that CHIP was associated with a nearly three-fold higher risk of NSCLC in non-smoking individuals living in areas with above-median PM_2.5_ levels (N = 109,635 individuals; HR 2.70, 95% CI: 1.60– 4.55, p = 0.0002; **Figure 1C**), while those living in below-median PM_2.5_ levels (N = 117,498 individuals) had no significant increased risk of CHIP-associated NSCLC (HR 1.45, 95% CI: 0.74–2.85, p = 0.28). The magnitude of interaction with PM_2.5_ was more pronounced for large CHIP clones (HR for interaction 1.72, 95% CI: 1.15–2.58, p-interaction = 0.009) and non-*DNMT3A* CHIP (HR for interaction 1.84, 95% CI: 1.06–3.22, p-interaction = 0.03). In contrast to PM_2.5_, coarse particulate air pollution levels (diameter 2.5-10µm) were not associated with NSCLC, and did not interact with CHIP status (correlation coefficient between PM_2.5_ and PM_2.5-10_: 0.22).

We then assessed whether PM_2.5_ levels were associated with increased CHIP prevalence. Consistent with previous observations in TOPMed cohorts^29^, we found no increase in CHIP prevalence with higher PM_2.5_ levels in unadjusted analyses, analyses adjusted for clinical covariates including smoking, nor in analyses stratified by smoking status (**Figure 2A**). When common CHIP driver genes were examined individually, *PPM1D*-CHIP was associated with PM_2.5_ levels, but only among current/former smokers (**Figure 2B**). Additionally, there was no correlation between CHIP variant allele fraction and PM_2.5_ levels, neither in unadjusted nor adjusted analyses (p = 0.81 and p = 0.84, respectively) nor when stratified by smoking status (**Figure 2C**). Altogether, these findings suggest that CHIP prevalence is not correlated with ambient air pollution levels.

**Figure 2.**
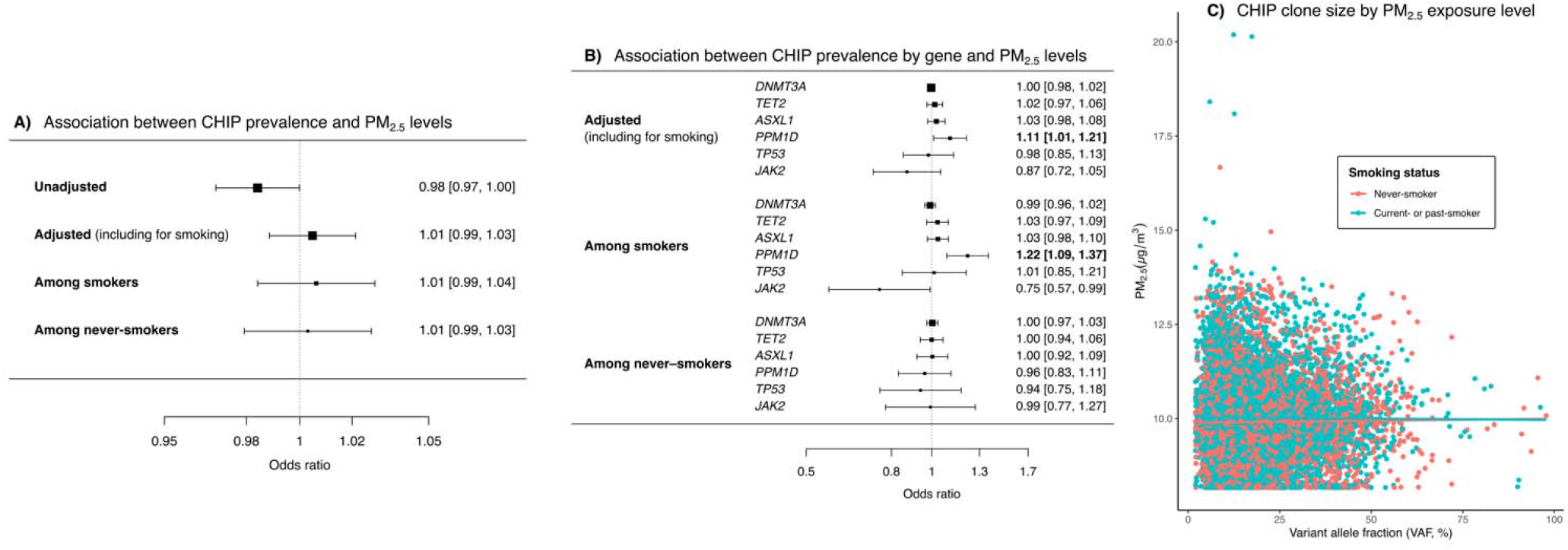
**A)** CHIP prevalence does not correlate with PM_2.5_ levels in models accounting for smoking status. **B)** *PPM1D*-CHIP is the only major CHIP subtype that correlates with PM_2.5_ levels, and the association was observed only among individuals who have smoked. In these analyses, CHIP as a binary trait is the independent variable and associations are therefore reported as odds ratios. **C)** CHIP clone size, as estimated by the variant allele fraction, is not associated with PM_2.5_ levels irrespective of smoking status.

Finally, we looked at circulating inflammatory markers with respect to CHIP status and PM_2.5_. CHIP, non-*DNMT3A* CHIP, large CHIP clones, and PM_2.5_ were each independently associated with greater baseline C-reactive peptide (CRP) levels in analyses adjusted for covariates including smoking (full list in *Methods*). We identified a significant positive interaction between CHIP status and PM_2.5_ levels with respect to their association with CRP levels (p-interaction = 0.01). Prior work has suggested that PM_2.5_ specifically activates IL-6 and IL-1β in the lung microenvironment.^4^ We posited that the circulating microenvironment may be a surrogate for the lung microenvironment. Serum IL-6 and IL-1β protein levels were available in a subset of individuals (∼10% of the cohort). We identified a significant positive interaction between CHIP × PM_2.5_ with respect to IL-1β levels (β 0.031 ± 0.015, p = 0.047) and IL-6 levels (β 0.06 ± 0.02, p = 0.003). The IL-1β association was attenuated after additional adjustment for potential batch effects (β 0.023 ± 0.015, p = 0.13) while the IL-6 association remained significant (β 0.06 ± 0.02, p = 0.002).

## Discussion

Between 10 and 25% cases of lung cancer worldwide are reported to occur in individuals who have never smoked.^30^ In the present work, we identify that 15% of NSCLC cases occur in never-smokers in the UK Biobank, and that small particulate matter air pollution (PM_2.5_) potentiates CHIP-associated NSCLC risk in this group. We observe that PM_2.5_ on its own is not associated with increased risk of NSCLC among never-smokers in the UKB; rather, it intensifies the association of CHIP and NSCLC in a gene × environment interaction. Based on prior work, the mechanism for this interaction is presumed to be increased inflammatory burden in the tumor microenvironment; concordant with this, we observed that CHIP and high PM_2.5_ levels are associated with more systemic inflammation when present together than either alone.

Our work was conducted in individuals residing in the UK, where PM_2.5_ ambient levels are among the lowest in the world (median: 9.93 µg/m^3^, maximum: 21.31 µg/m^3^), whereas many countries see upwards of 5-7× higher average levels (**Supplemental Figure 1**) with concordantly higher rates of lung cancer among non-smokers.^4^ Despite the lower levels and a narrower range of PM_2.5_ levels in our dataset, we observed a gene × environment interaction with CHIP. CHIP was not more prevalent in individuals exposed to greater levels of air pollution within our dataset, in line with previous observations.^29^ As such, we do not necessarily expect CHIP to be more prevalent in countries with higher levels of air pollution. However, CHIP is a common condition across populations (on average affecting >10% of individuals aged 65 and older); and so, the contribution of CHIP to global air pollution-related lung cancer is expected to be significant and could inform future screening strategies.

Limitations of our work include that PM_2.5_ exposure levels were estimated for each person based on their reported place of residence at enrollment using Land Use Regression (LUR) estimates^26^ for the year 2010 (approximately mid-way through the UK Biobank enrollment period). This single-year estimate may not accurately reflect their lifetime PM_2.5_ exposure, particularly if they have moved within or immigrated to the UK. Additionally, smoking status is self-reported in the UK Biobank, which may underestimate smoking prevalence.^31^ Finally, we were not able to directly assess our mechanistic hypothesis linking CHIP and NSCLC (*i*.*e*., that CHIP-mutated proinflammatory macrophages would promote the clonal expansion of lung tissue with existing cancer-driver mutations).

## Data availability

CHIP calls for UKB participants have been returned to the UKB Access Management System (AMS) and will be available to all registered researchers once processed by the UKB AMS team.^22^ All other data is available to registered UKB researchers.

## Funding

This work was supported by NIH grants DP5 OD029586, R01 AG083736, a Burroughs Wellcome Fund Career Award for Medical Scientists, the E.P. Evans Foundation, RUNX1 Research Program, a Pew-Stewart Scholar for Cancer Research award, supported by the Pew Charitable Trusts and the Alexander and Margaret Stewart Trust to A.G.B., a National Institutes of Health/National Cancer Institute (NIH/NCI) grant #U01CA253560 awarded to M.C.A., a Canadian Institute of Health Research (CIHR) Project Grant (#202010PJT-451137) awarded to M.J.R, a PSI Foundation grant #2020-1910 awarded to C.V. and M.J.R, and a CIHR Canadian Graduate Scholarship (CGS) – Doctoral awarded to C.V.

**Supplemental Figure 1.**
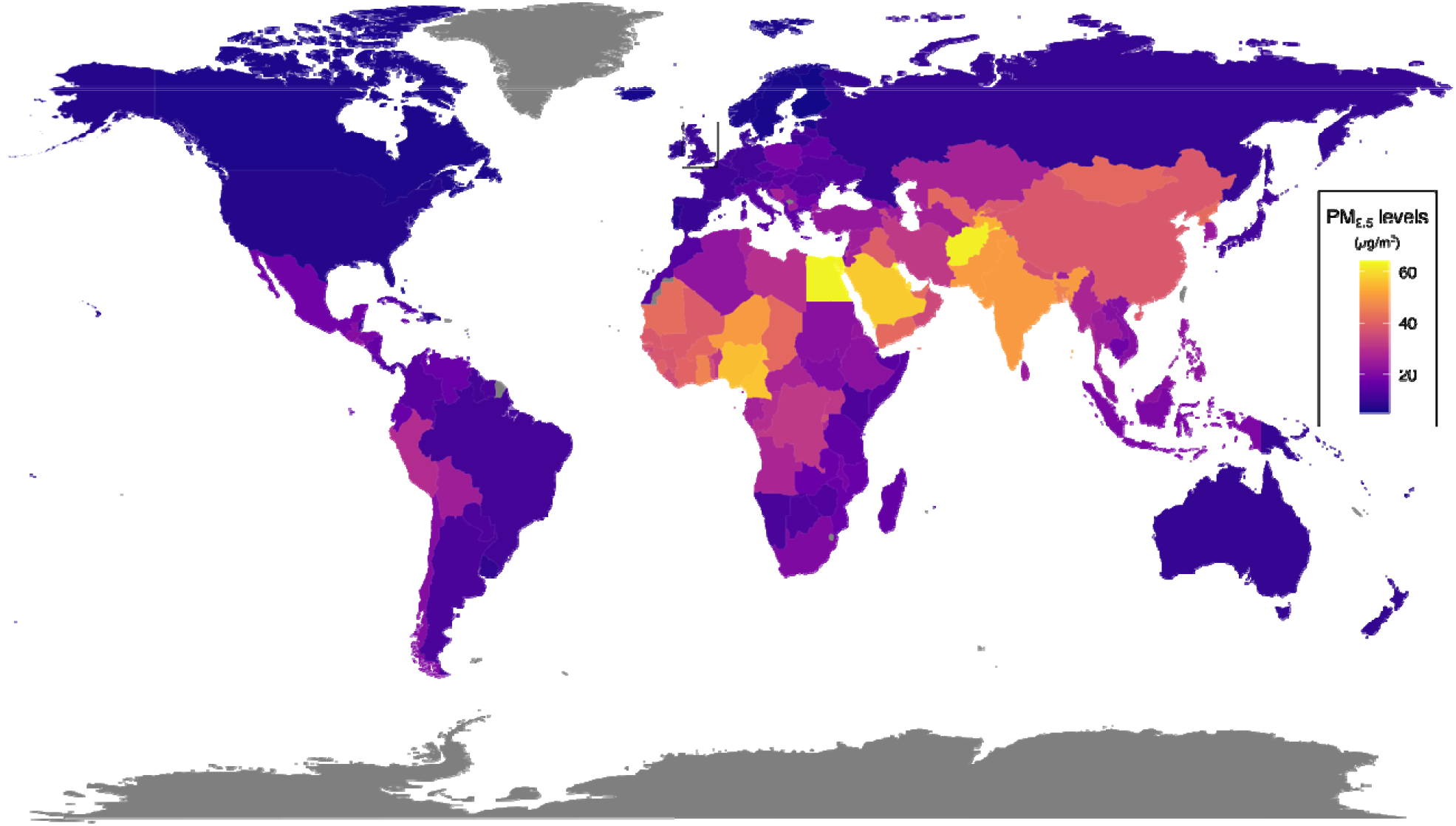
Average concentration of small particulate matter air pollution (PM_2.5_) by country. Data from the World Health Organization (WHO) Air pollution data portal (https://www.who.int/data/gho/data/themes/air-pollution/modelled-exposure-of-pm-air-pollution-exposure). The area included in this study (UK) is highlighted with a box.

