## Supplemental Figure 1 for "Clonal hematopoiesis of indeterminate potential-associated non-small cell lung cancer risk is potentiated by small particulate matter air pollution among non-smokers: a novel somatic variant–environment interaction"

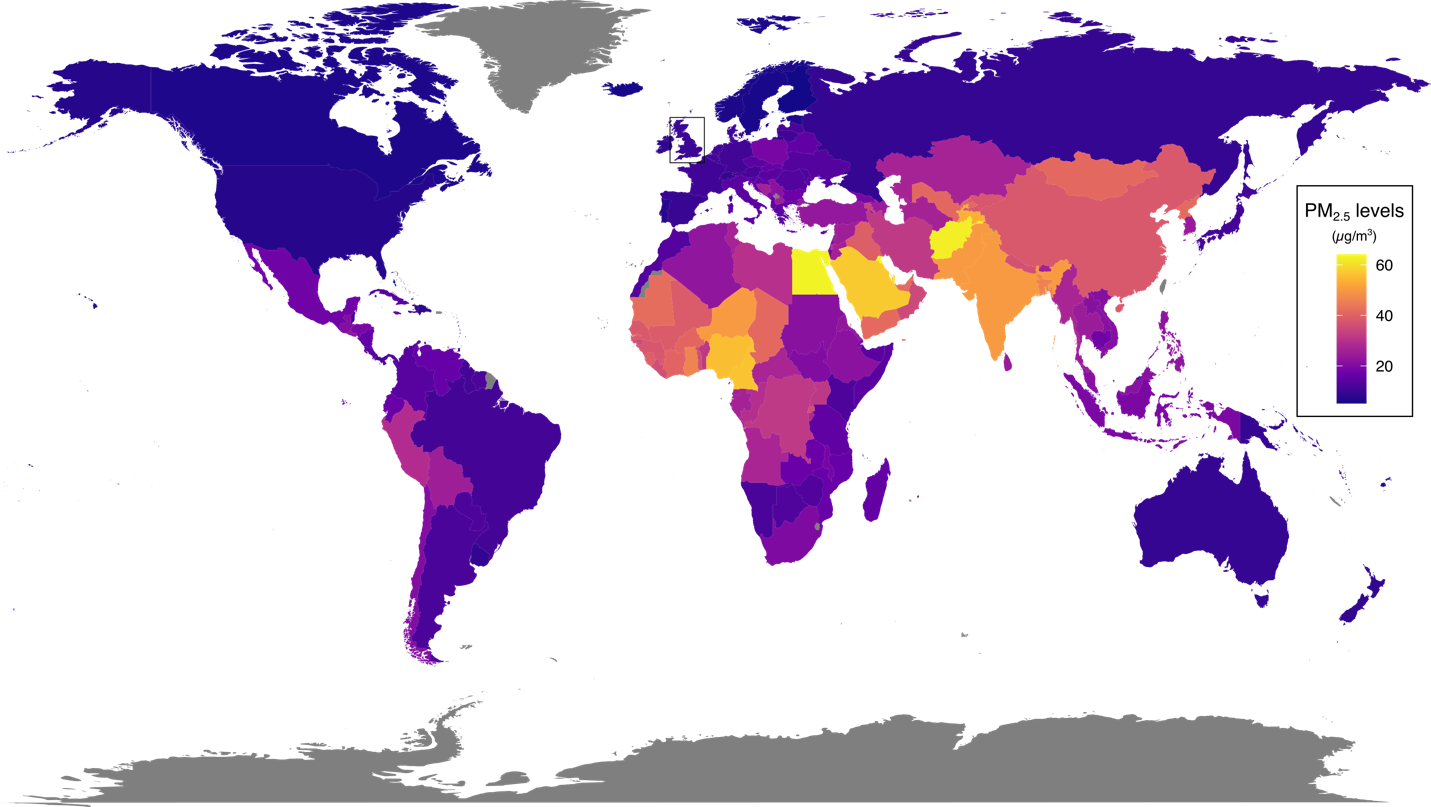

**Supplemental Figure 1**. Average concentration of small particulate matter air pollution (PM_2.5_) by country. Data from the World Health Organization (WHO) Air pollution data portal (<https://www.who.int/data/gho/data/themes/air-pollution/modelled-exposure-of-pm-air-pollution-exposure>). The area included in this study (UK) is highlighted with a box.
